# Associations of Racial and Ethnic Composition of School Districts with Wildfire Smoke Exposure and Reduced In-Person Learning Among Schoolchildren During the Pandemic in the United States

**DOI:** 10.1101/2023.09.25.23296000

**Authors:** Daniel Kim

## Abstract

**Introduction:** The White House has called for an “all-hands-on-deck” response to address chronic absenteeism and disrupted learning among K-12 students that spiked during the COVID-19 pandemic due to school closures and shifts to remote learning in the United States. Such learning disruptions are linked to test score declines with lifelong implications for earnings and well-being. This study aimed to identify higher-risk student populations that might benefit from interventions to reduce such disruptions and related inequities by estimating the linkages between the racial and ethnic composition of U.S. K-12 school districts as predictors of average wildfire smoke levels and year-over-year declines in in-person student visits to schools during the pandemic.

**Methods:** Using multivariable logistic regression, 5-year (2012-2016) mean percentages of non-White students grouped into quartiles were investigated as predictors of school district-level outcomes consisting of: 1) 5-year (2012-2016) mean levels of wildfire smoke fine particulate matter 2.5 μm or smaller in diameter (PM_2.5_) above 35 μg/m^3^ (EPA health-based standard) during school days for grade 3-8 students from August 15 to June 15 of the following year; and 2) being above the median for the mean decline in in-person student attendance (vs. the same month in 2019), from September 2020 to May 2021 and September 2021 to May 2022.

**Results:** Across 11,190 (82.4% of all) U.S. school districts, the highest (vs. lowest) quartile for the percentage of non-White students predicted a nearly 3-fold higher odds (adjusted odds ratio, AOR=2.82; 95% CI=2.14-3.72; *P*<.001) for high wildfire smoke exposure and a 9-fold higher odds (AOR=9.12; 95% CI=7.15-11.63; *P*<.001) for substantially reduced in-person learning levels. Successively higher odds were observed in higher quartiles (*P* for trend <.001). Similar patterns were seen when high (vs. low) percentages of Asian-, Black-, and Hispanic-American students were modeled simultaneously.

**Conclusions:** U.S. school districts with higher percentages of non-White students showed a convergence of elevated odds of wildfire smoke exposure and distance learning consistent with dose-response relationships. These concomitant risks could be mitigated by school-based air filtration interventions that reduce wildfire smoke PM_2.5_ and transmission of airborne particulates carrying the coronavirus. One potential cost-effective intervention to address these joint risks would be the placement of Corsi-Rosenthal boxes (DIY inexpensive portable air filtration devices shown to effectively reduce PM_2.5_ concentrations) in school classrooms and other school indoor areas. Supplying Corsi-Rosenthal boxes to school districts could markedly reduce learning disruptions and wildfire smoke and COVID-19 health impacts and their related inequities at a cost of less than $1 billion annually, 0.5% of the $190 billion in federal relief available to school districts to address pandemic-related needs including learning loss.

## Introduction

The White House has called for an “all-hands-on-deck” response to address chronic absenteeism and disrupted learning among K-12 students that spiked during the COVID-19 pandemic due to school closures and shifts to remote learning.^1^ Such learning disruptions are linked to test score declines with lifelong implications for earnings and well-being.^2^ Exposure to wildfire smoke during school days further predicts lower test scores among schoolchildren^3^ and can exacerbate clinical conditions in children such as asthma that contribute to absenteeism.

Average wildfire smoke exposure levels will likely increase over time nationwide due to global warming, and while the coronavirus pandemic has been declared over, COVID-19 school outbreaks which disproportionately affected communities of color are ongoing. This study aimed to identify higher-risk student populations that might benefit from interventions to reduce learning disruptions and related inequities by estimating the linkages between the racial and ethnic composition of school districts as predictors of average wildfire smoke levels and year-over-year declines in in-person student visits to schools during the pandemic.

## Methods

Using multivariable logistic regression, 5-year (2012-2016) mean percentages of non-White students grouped into quartiles were investigated as predictors of school district-level outcomes consisting of: 1) 5-year (2012-2016) mean levels of wildfire smoke fine particulate matter 2.5 μm or smaller in diameter (PM_2.5_) above 35 μg/m^3^ (EPA health-based standard) during school days for grade 3-8 students from August 15 to June 15 of the following year^3^; and 2) being above the median for the mean decline in in-person student attendance (vs. the same month in 2019), from September 2020 to May 2021 and September 2021 to May 2022, derived from a public database that tracked month-to-month K-12 distance learning for public schools using aggregated, anonymized mobile phone GPS data.^4^

All models controlled for the mean percentage of students eligible for free/reduced-price school lunches (as a proxy for socioeconomic disadvantage), total student enrolment, number of schools, and state fixed effects.

Standard errors were adjusted for clustering of school districts within states. In supplementary analyses, high (vs. low) percentages of Black-, Hispanic-, and Asian-American students categorized using median values were examined.

## Results

Across 11,190 (82.4% of all) U.S. school districts, the highest (vs. lowest) quartile for the percentage of non-White students predicted a nearly 3-fold higher odds (adjusted odds ratio, AOR=2.82; 95% CI=2.14-3.72; *P*<.001) for high wildfire smoke exposure (Fig. 1) and a 9-fold higher odds (AOR=9.12; 95% CI=7.15-11.63; *P*<.001) for substantially reduced in-person learning levels (Fig. 2). Successively higher odds were observed in higher quartiles (*P* for trend <.001). Similar patterns were seen when high (vs. low) percentages of Asian-, Black-, and Hispanic-American students were modeled simultaneously (Figs. 3 and 4).

**Figure 1.**
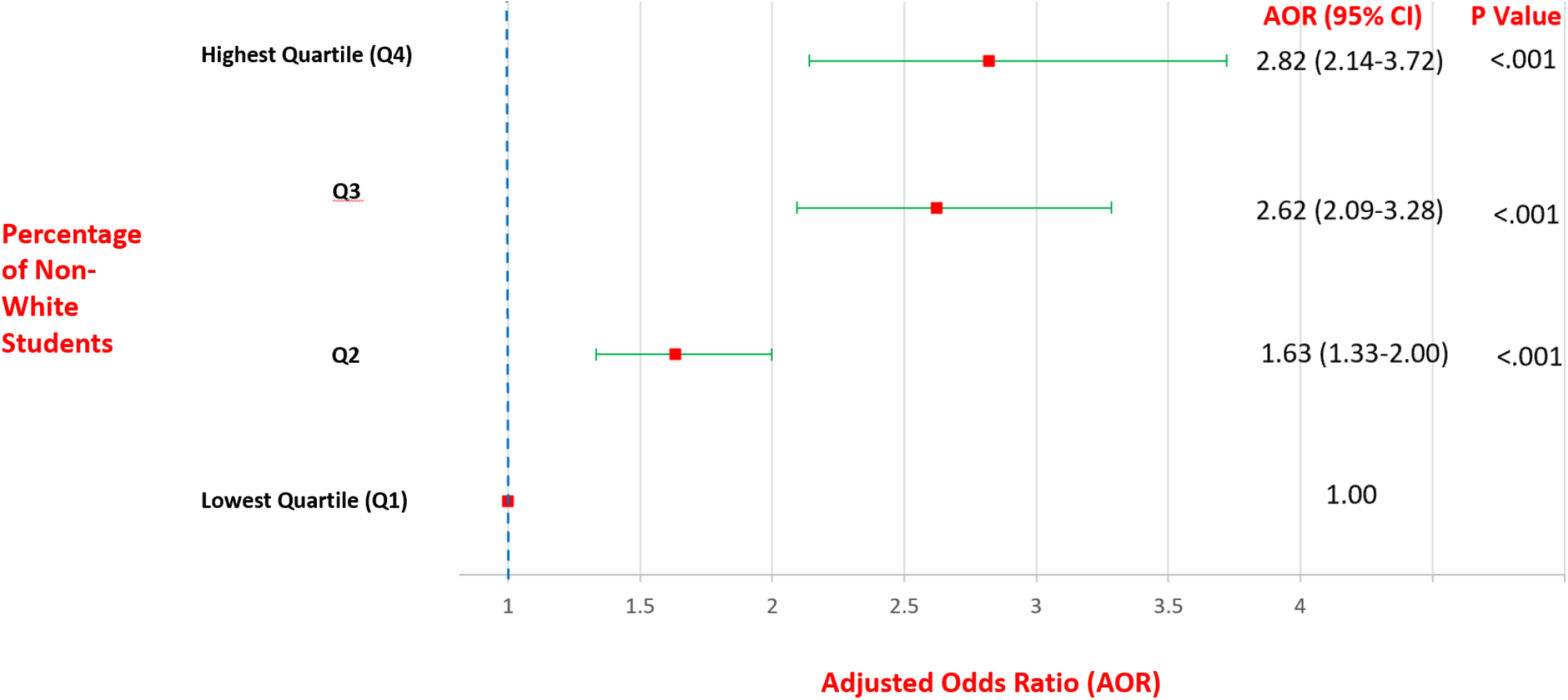
Multivariable-adjusted odds ratios of mean levels of wildfire smoke PM_2.5_ above 35 μg/m^3^ during school days for grade 3-8 students according to percentage of non-White students across 11,190 school districts in the United States, 2012-2016. All models are adjusted for the mean percentage of students eligible for free/reduced-price school lunches, total student enrolment, number of schools, and state fixed effects.

**Figure 2.**
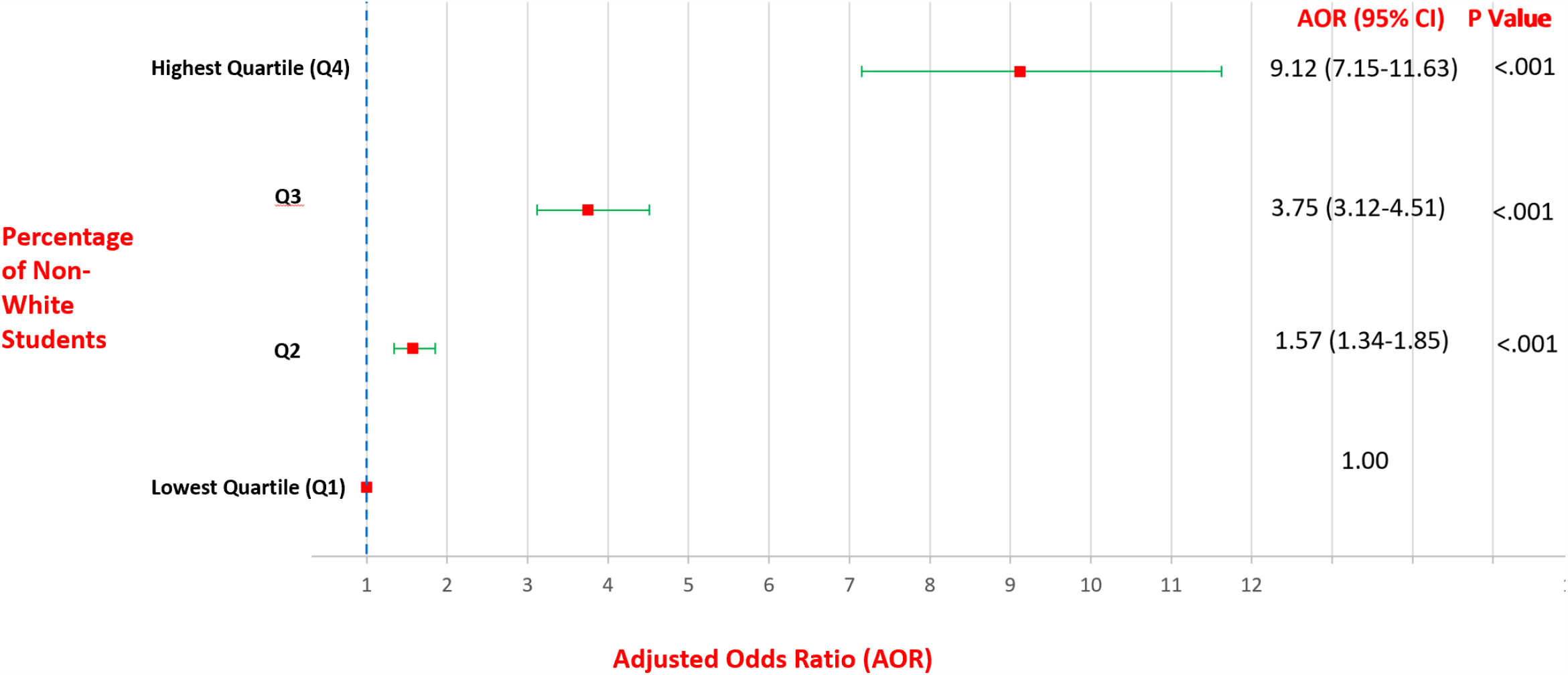
Multivariable-adjusted odds ratios of high mean decline in in-person student attendance from September 2020 to May 2021 and September 2021 to May 2022 (vs. same month in 2019) for K-12 students according to percentage of non-White students across 11,190 school districts in the United States, 2012-2016. All models are adjusted for the mean percentage of students eligible for free/reduced-price school lunches, total student enrolment, number of schools, and state fixed effects.

**Figure 3.**
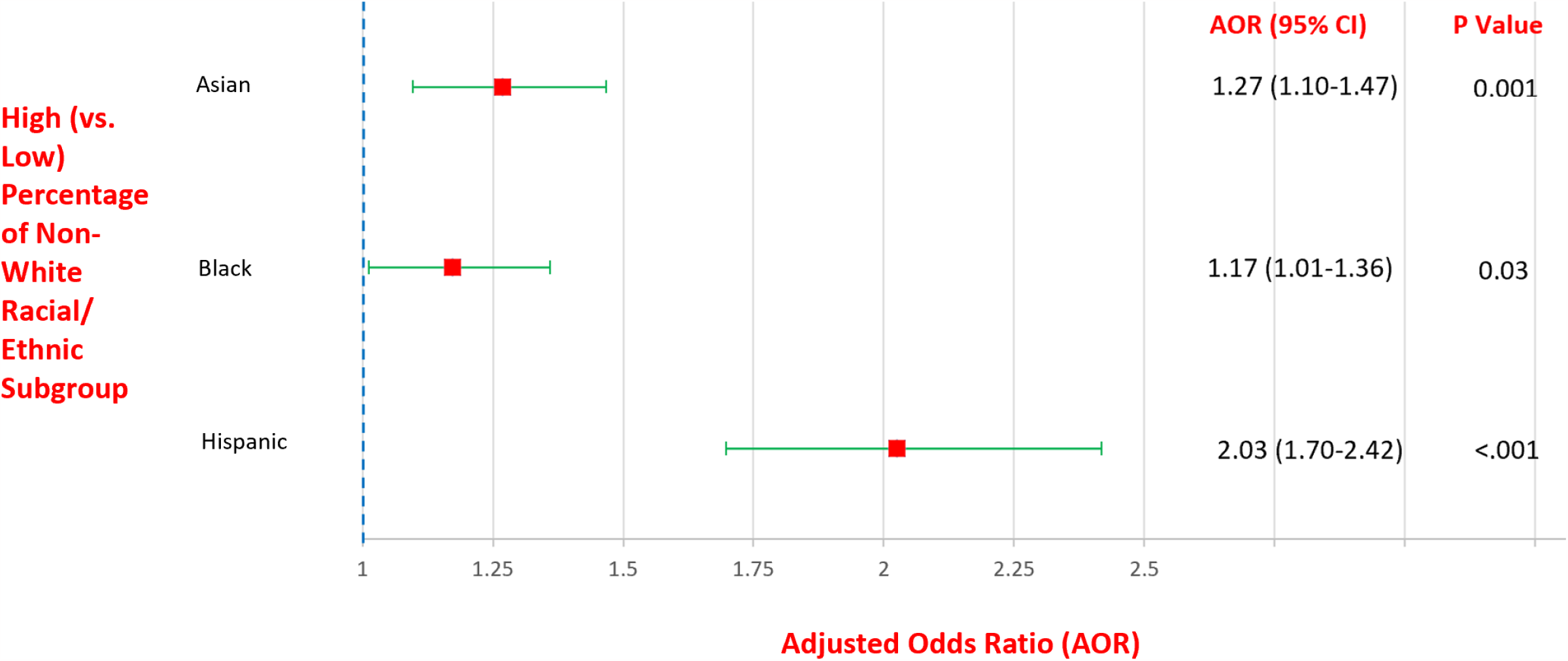
Multivariable-adjusted odds ratios of mean levels of wildfire smoke PM_2.5_ above 35 μg/m^3^ during school days for grade 3-8 students according to high (vs. low) percentage of Asian-, Black-, and Hispanic-American students across 11,190 school districts in the United States, 2012-2016. All models are adjusted for the mean percentage of students eligible for free/reduced-price school lunches, total student enrolment, number of schools, and state fixed effects.

**Figure 4.**
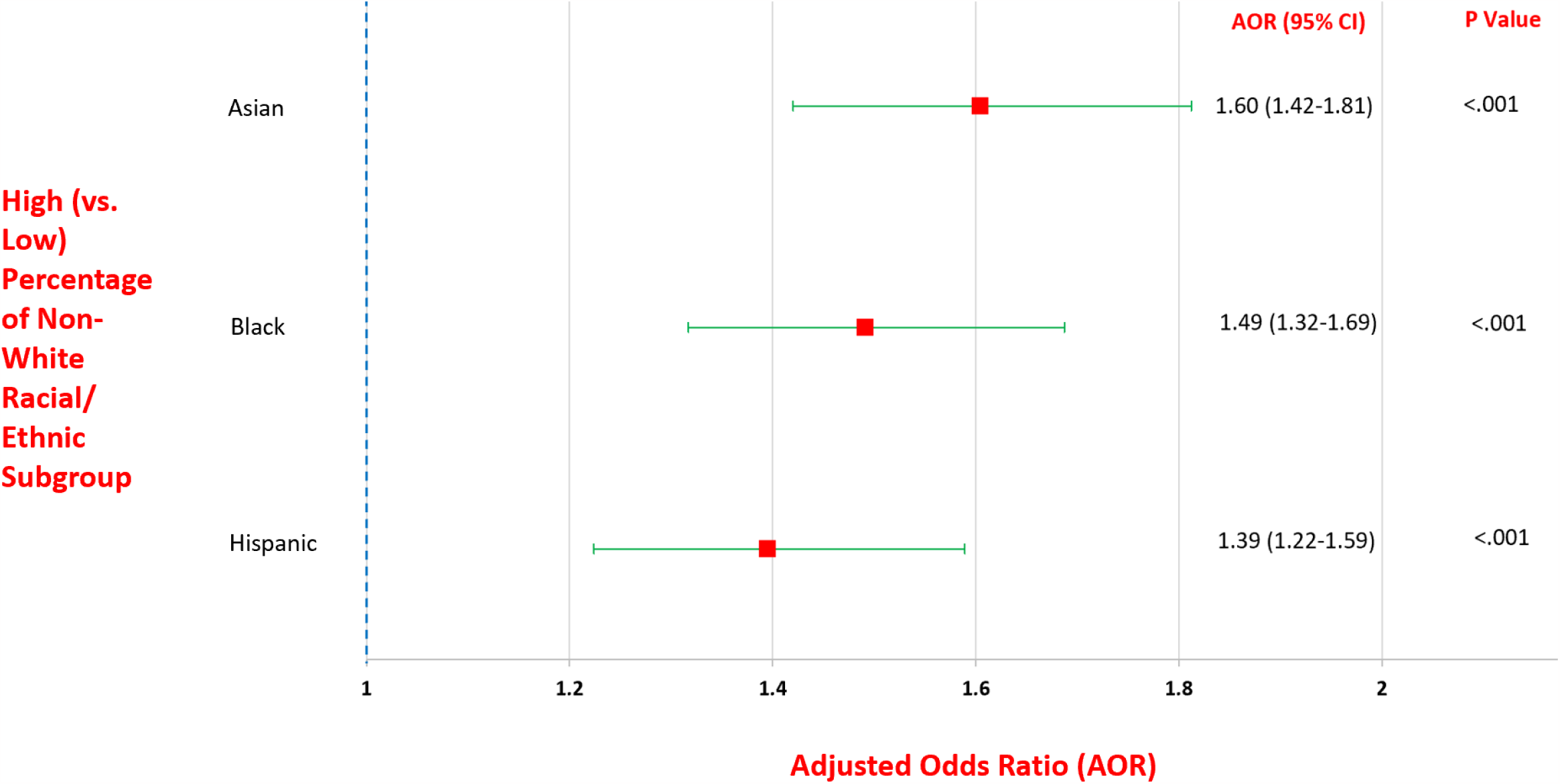
Multivariable-adjusted odds ratios of high mean decline in in-person student attendance from September 2020 to May 2021 and September 2021 to May 2022 (vs. same month in 2019) for K-12 students high (vs. low) percentage of Asian, Black-, and Hispanic-American students across 11,190 school districts in the United States, 2012-2016. All models are adjusted for the mean percentage of students eligible for free/reduced-price school lunches, total student enrolment, number of schools, and state fixed effects.

## Discussion

U.S. school districts with higher percentages of non-White students showed a convergence of elevated odds of wildfire smoke exposure and distance learning consistent with dose-response relationships. These concomitant risks could be mitigated by school-based air filtration interventions that reduce wildfire smoke PM_2.5_ and transmission of airborne particulates carrying the coronavirus.

One potential cost-effective intervention to address these joint risks would be the placement of Corsi-Rosenthal boxes (DIY inexpensive portable air filtration devices shown to effectively reduce PM_2.5_ concentrations) in school classrooms and other school indoor areas.^5^ Supplying Corsi-Rosenthal boxes to school districts belonging to the highest three quartiles of percentages of non-White students nationally could markedly reduce learning disruptions and wildfire smoke and COVID-19 health impacts and their related inequities at less than $1 billion annually (Table S1 in Appendix). This total cost represents just 0.5% of the $190 billion in federal relief available to school districts until September 2024 to address pandemic-related needs including learning loss.^6^

A key strength of this analysis over prior studies is its co-adjustment for socioeconomic disadvantage and racial/ethnic composition in regression models, thereby reducing confounding. Study limitations include its measurement of racial and ethnic composition and wildfire smoke exposure during pre-pandemic time periods;^3^ its reliance on GPS data as a proxy for personal location of schoolchildren; and its ecological design.^4^

## Data Availability

All data produced in the present study are available upon reasonable request to the author.

### Appendix

**Table S1.**
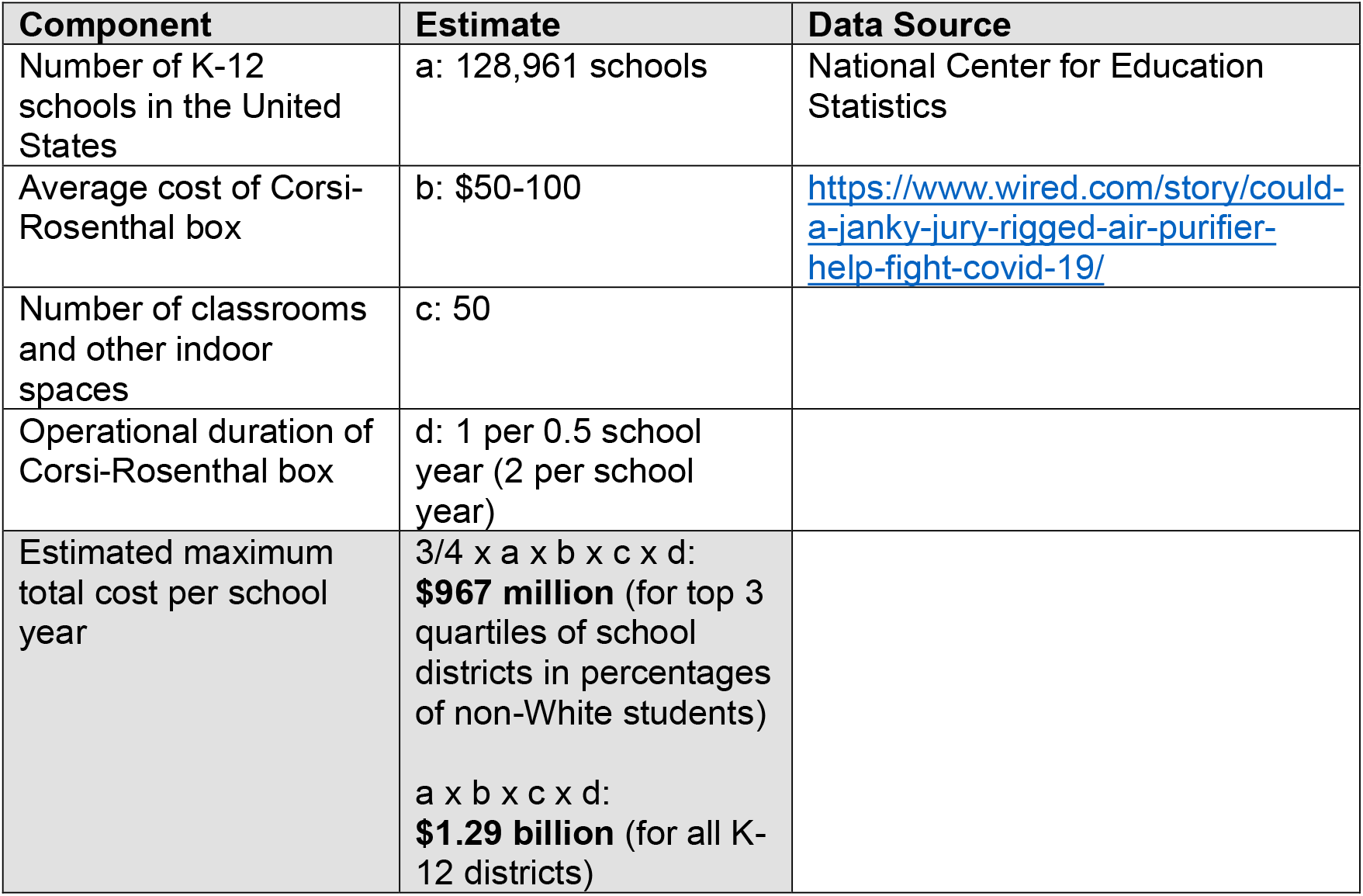
Estimated maximum total cost of supplying Corsi-Rosenthal boxes to K-12 school districts in the United States per school year.

| Component | Estimate | Data Source |
| --- | --- | --- |
| Number of K-12 schools in the United States | a: 128,961 schools | National Center for Education Statistics |
| Average cost of Corsi-Rosenthal box | b: \$50-100 | https://www.wired.com/story/could-a-janky-jury-rigged-air-purifier-help-fight-covid-19/ |
| Number of classrooms and other indoor spaces | c: 50 |  |
| Operational duration of Corsi-Rosenthal box | d: 1 per 0.5 school year (2 per school year) |  |
| Estimated maximum total cost per school year | $\frac{3}{4} \times a \times b \times c \times d$:<br>**\$967 million** (for top 3 quartiles of school districts in percentages of non-White students)<br>$a \times b \times c \times d$:<br>**\$1.29 billion** (for all K-12 districts) | |

